# Evolution of Human Respiratory Virus Epidemics

**DOI:** 10.1101/2020.11.23.20237503

**Authors:** Nash D. Rochman, Yuri I. Wolf, Eugene V. Koonin

## Abstract

**Background:** While pathogens often evolve towards reduced virulence, many counterexamples are evident. When faced with a new pathogen, such as SARS-CoV-2, it is highly desirable to be able to forecast the case fatality rate (CFR) into the future. Considerable effort has been invested towards the development of a mathematical framework for predicting virulence evolution. Although these approaches accurately recapitulate some complex outcomes, most rely on an assumed trade-off between mortality and infectivity. It is often impractical to empirically validate this constraint for human pathogens.

**Results:** Using a compartment model with parameters tuning the degree to which symptomatic individuals are isolated and the duration of immunity, we reveal kinetic constraints where the variation of multiple parameters in concert leads to decreased virulence and increased pathogen fitness, whereas independent variation of the parameters decreases pathogen fitness. Smallpox, SARS-CoV-2, and Influenza are analyzed as diverse representatives of human respiratory viruses. We show that highly virulent viruses, such as Smallpox, are likely often constrained by host behavior, whereas moderately virulent viruses, such as SARS-CoV-2, appear to be typically constrained by the relationship between the duration of immunity and CFR.

**Conclusions:** The evolution of human respiratory epidemics appears to be often kinetically constrained and a reduction in virulence should not be assumed. Our findings imply that, without continued public health intervention, SARS-CoV-2 is likely to continue presenting a substantial disease burden. The existence of a parameter regime admitting endemic equilibrium suggests that herd immunity is unachievable. However, we demonstrate that even partial isolation of symptomatic individuals can have a major effect not only by reducing the number of fatalities in the short term but also by potentially changing the evolutionary trajectory of the virus towards reduced virulence.

## Background

The rates of morbidity, mortality, and infection determine whether a pathogen is tolerated by its host and, in turn, the survival of the pathogen itself. Constraints imposed by the host behavior and pathogen biology prevent independent variation of these rates. Therefore, trends in virulence evolution can be predicted only through understanding these constraints. Although comprehensive models of the evolution of virulence capable of describing complex environments have been developed[1-5], most studies to date impose constraints assumed from first principles and lacking experimental or empirical validation[6,7]. Most commonly, a trade-off function is assumed[8,9] between the rate at which the pathogen is transmitted between hosts and the case fatality rate (CFR).

Hosts with high pathogen loads are more likely to die than those with lower loads, but they also shed the pathogen at increased rates and therefore could transmit it to a greater number of new hosts. However, this straightforward picture is complicated by a landscape of opposite evolutionary outcomes. For example, leaky vaccination has been suggested to increase virulence in malaria [10,11] and decrease virulence for diphtheria and pertussis [12], likely due to differences in the cost of toxin production [13]. This demonstrates how predictions of specific outcomes useful for informing public health intervention cannot be generalized across pathogens. The infectivity-virulence trade-off has been demonstrated for malaria[11], but not for most human pathogens and is extremely hard to validate empirically due to the impracticality of comprehensive contact tracing. Thus, models that avoid the use of this constraint as a parameter have the potential to produce useful observations and predictions.

Towards this goal, we explore a range of epidemiological outcomes for human pathogens modelled using available data on well-characterized respiratory viruses. The apparent inverse relationship between the time during which the host is asymptomatic but infectious and the mortality rate[14-22], as well as the difficulty of vaccination against lower mortality viruses, such as Influenza, relative to higher mortality viruses, such as Smallpox, imply the existence of intrinsic constraints that could link the duration of immunity and mortality. We demonstrate that, in addition to trade-offs, virulence evolution is often subject to “kinetic constraints” that prevent virulence reduction by imposing a barrier in the fitness landscape, analogous to an energetically favorable chemical reaction with a high activation energy. Virulence evolution would be thermodynamically constrained if it were impossible to both increase pathogen fitness and decrease CFR. In contrast, virulence evolution is kinetically constrained when there exists a parameter regime where pathogen fitness is higher and CFR is lower, but accessing this regime requires simultaneously modifying multiple parameters, some of which might be determined by host behavior. In other words, there is a path to decreased CFR on the fitness landscape, but it is narrow. We show that, for high CFR viruses such as Smallpox, the relationship between transmission rate and mortality is likely a kinetic constraint, rather than an actual trade-off, whereas for intermediate CFR viruses, such as SARS-CoV-2, there is a kinetic constraint between immunity and mortality. Analysis of such constraints could open avenues for prediction of epidemic outcomes and quantitative validation of such predictions.

## Methods

The introduction of a novel pathogen into a host population is likely to result in one of three outcomes. 1) The number of fatalities is large enough for the pathogen to wipe out the host population (as a result, the pathogen itself also goes extinct) (Fig. 1A). 2) The number of infections is large enough, whereas the number of fatalities is small enough, such that the pathogen creates a bottleneck in the susceptible host population. With all hosts either infected or immune, the pathogen is eliminated from the host population (Fig. 1B). 3) The number of infections is small enough such that a bottleneck in the susceptible population is avoided and long-term co-evolution with the host is possible if the number of infections is not too small and the basic reproduction number is greater than one, *R*_*0*_>1 (Fig. 1C). With *R*_*0*_>1, a state of stable endemic equilibrium can be reached where the fraction of the host population susceptible to infection remains constant (up to fluctuations). However, if the pathogen-associated fatality exceeds the birth-rate of the host population, the host-pathogen relationship becomes unsustainable in the long term. Such an unsustainable relationship would pose an intense selective pressure on the host population, likely resulting in the extinction of the pathogen through the modification of host behavior or the emergence of resistant hosts.

**Figure 1:**
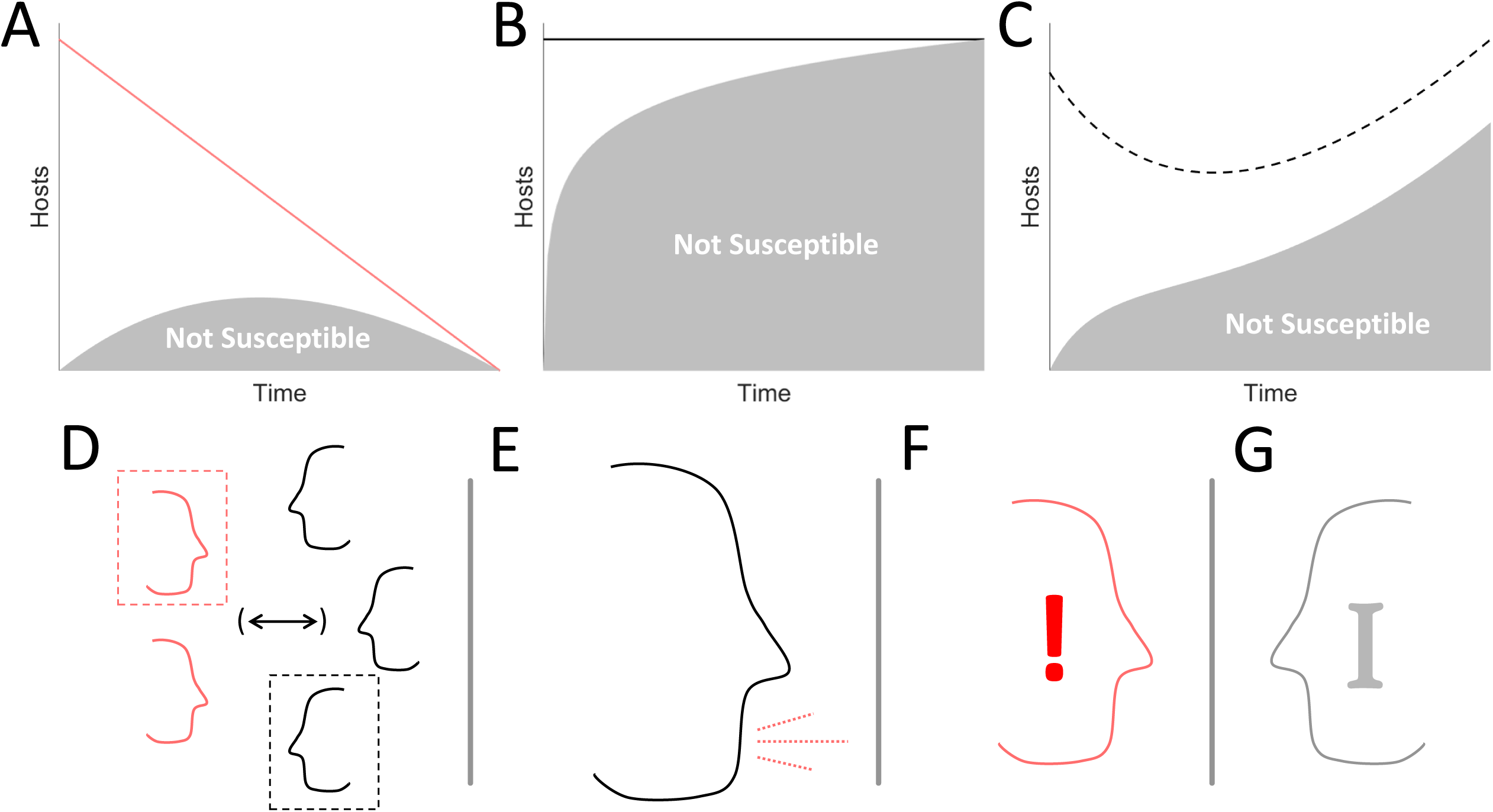
Key factors which determine epidemiological outcomes. **A-C**. Lines indicate total number of hosts, shaded areas indicate the fraction of hosts which are not susceptible. **A**. Host extinction. **B**. A bottleneck in the susceptible population. **C**. A balance between infections, mortality, and birthrate **D**. Host interaction, quarantine, and isolation. **E**. Infectivity. **F**. Virulence/mortality. **G**. Immunity.

Which of these courses an epidemic follows, largely depends on the balance of four factors. 1) The frequency of host-host interaction, with or without isolation of infected individuals or prophylactic quarantine (Fig. 1D). 2) The infectivity of the pathogen, that is, the likelihood of an uninfected host to become infected after interacting with an infected host (Fig. 1E). 3) The virulence of the pathogen, that is, the likelihood that an infected host will experience symptoms or die (Fig. 1F). 4) The duration of host immunity post infection (Fig. 1G). Although the effect of tuning each of these parameters often appears obvious - for example, decreasing the frequency of host-host interaction almost always decreases the number of infections - many counterintuitive observations become apparent. For example, decreasing the frequency of host-host interaction under conditions that would otherwise lead to a bottleneck in the susceptible population (Fig. 1B) can result in stable co-evolution with the host which, avoiding pathogen extinction, ultimately increases the total number of infections sustained over time. In an effort to better delineate how the host-pathogen relationship varies across the space of these factors, we constructed the following model.

Hosts are assigned one of the four possible states: 1) immune *I*, 2) susceptible *S*, 3) asymptomatic *A*, and 4) symptomatic or “clinical” *C*. New hosts are assumed to be born susceptible at a rate *k*_*B*_ and a baseline death rate *k*_*D*_ is assumed constant across all compartments. Susceptible hosts can be infected by coming into contact with either asymptomatic or clinical hosts. Asymptomatic hosts either recover at rate *k*_*R*_ or progress to the clinical compartment at rate *k*_*P*_. Clinical hosts either recover at rate *k*_*R*_ or die due to the pathogen at rate *k*_*DV*_. Recovery confers immunity which is then lost at rate *k*_*L*_. At endemic equilibrium, the parameter 0 ≤ α ≤ 1 can be used to represent immunity such that *k*_*L*_ = (1 − α)*k*_*R*_(*A* + *C*)/*I*. At the beginning of the epidemic, α ≈ 1. The population is well mixed, with the exception of the clinical compartment, a fraction (1 − *β*) of which is isolated and cannot infect susceptible hosts. The rate at which susceptible hosts become infected is the triple product of the rate of contact between hosts, the fraction of the population (not isolated) which is infected, and the probability of infection upon contact: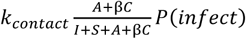. For simplicity, we consider the product, *k*_*I*_ ≡ *k*_*contact*_*P*(*infect*), which depends on both host behavior and pathogen biology. This yields the system of ordinary differential equations (Fig. 2A):

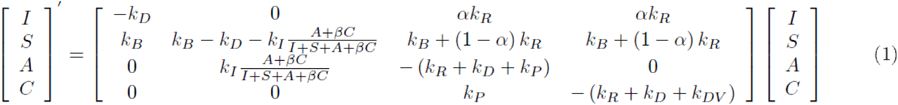

**Figure 2:**
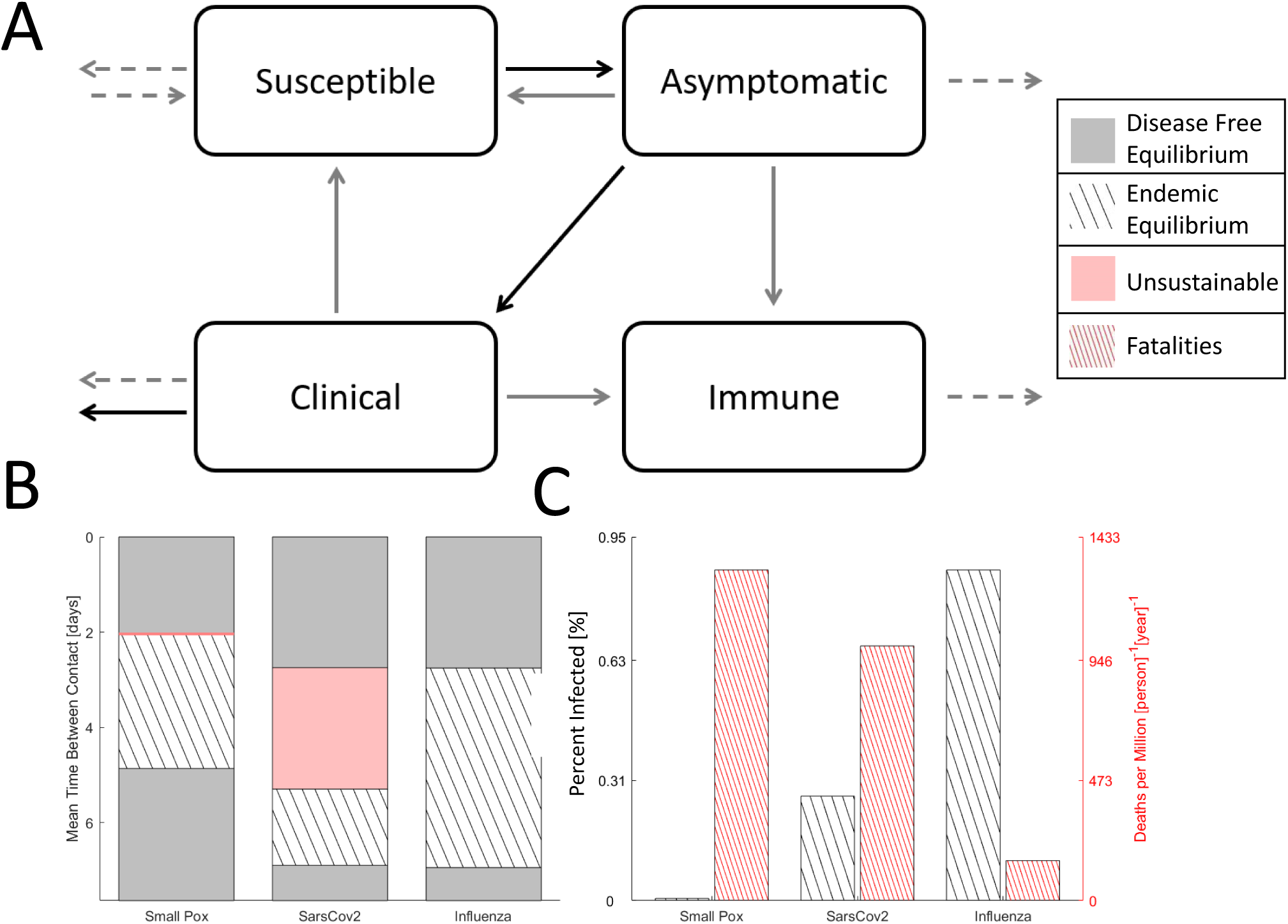
Four compartment model of an epidemic. **A**. Cartoon of the compartment model. **B**. Disease free equilibrium (gray), endemic equilibrium (striped), and host population decline (red) for three analyzed viruses over a range of host contact rates. The decline region for Smallpox is narrow and not shown to scale. **C**. For endemic equilibrium, the fraction of the host population infected and the death rate.

With two infected states (asymptomatic and clinical) and isolation, ISAC is a simple model within the range of models [23-25] that have been developed in response to the SARS epidemic.

The basic reproduction number, *R*_*0*_, for this model can be derived through the construction of next generation matrices[26] (see Appendix A in Additional File 1):

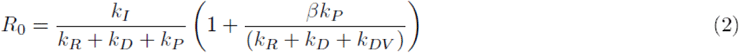

Short term dynamics are determined by the *R*_*0*_, value. When *R*_*0*_<1, the pathogen will go extinct. When *R*_*0*_>1, a wide range of dynamics are possible, depending on the parameter regime. In most cases, a unique, stable endemic equilibrium exists [27,28]; however, if the susceptible population first reaches a bottleneck (Fig. 1B), the pathogen could go extinct. At endemic equilibrium, the fraction of the total population, *N*, in each compartment, *X*, is constant: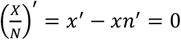. The case with the simplifying assumption of a constant total population is presented in Appendix B (Additional File 1). More generally, *n*^′^ = *k*_*B*_ − *k*_*D*_ − *ck*_*DV*_ which yields:

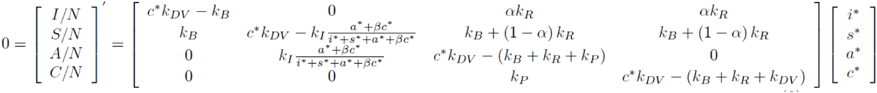

This system can be solved to yield a fourth order polynomial with respect to *c*_*_ (we used the MATLAB symbolic toolbox[29], see Appendix C in Additional File 1) which is cumbersome enough that a numerical solution appears preferable; however, endemic equilibrium requires a constant or growing population 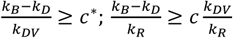. For human populations and pathogens, it is reasonable to assume that the birth rate is much lower than the recovery rate, 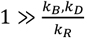, yielding the limit: 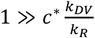.

Thus, for an endemic equilibrium to exist, either the clinical compartment has to be very small or the death rate due to the virus has to be very low compared to the recovery rate (in the opposite limit, when the birth rate is high, parameter regimes exist where neither endemic equilibrium is reached nor does either the host population or the virus go extinct). This allows us to linearize the model with respect to either the size of the clinical compartment, for pathogens with high mortality, or the ratio of the death rate to the recovery rate, for low mortality pathogens. For the high mortality case (see Appendix D in Additional File 1), the linearized model yields a unique, stable analytic solution for endemic equilibrium, given a sufficiently large *α* corresponding to at least partial immunity whenever *R*_0_ > 1[27,28]. In this case, the additional constraint *k*_*P*_ > *k*_*R*_ + *k*_*DV*_, which largely holds for the pathogens considered here, is applied for convenience. For the low mortality case (see Appendix E in Additional File 1), the stability of the solution depends on the parameters, and both stability and the general solution for the critical point are calculated numerically; however, analytic forms for endemic equilibria in the stricter limit 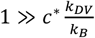 are provided.

The parameters were fit for three respiratory pathogenic viruses: Smallpox, SARS-CoV-2, and Influenza representing a range of phenotypes (Table 1). Here, *k*_*B*_ and *k*_*D*_ are fixed whereas *k*_*I*_ is varied. Then, *k*_*R*_ and *k*_*P*_ are fit to an estimated disease course for a host which is asymptomatic and infectious for the time *t*_*P*_ = 1/*k*_*P*_, and symptomatic and infectious for the time *t*_*R*_ = 1/*k*_*R*_ before recovering. For simplicity, death due to infection is assumed to occur only during the symptomatic and infectious phase: 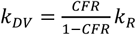. Smallpox is modelled with a CFR of 30%, mean time to recovery 1 week, and no asymptomatic and infectious period.

**Table 1:** Significant Human Respiratory Viruses.

| Name | Case Fatality Rate | Time Asym. N.Inf. (days) | Asym. Inf. | Time Sym. (days) |
| --- | --- | --- | --- | --- |
| MERS-CoV | 0.35 | 2--14 | minimal | up to 90 |
| Variola Major (Smallpox) | 0.3 | 7--19 | assumed no | 9--11 |
| SARS-CoV | 0.15 | 6 | assumed no | 20 |
| Measles Morbivirus | 0.04-0.1 | 6--21 | < one week | 6--8 |
| SARS-CoV-2 | <0.02 | 5--14 | < one week | 21 |
| Influenza | <0.002 | 2 | < one week | 3 |
| Mumps rubulavirus | near 0 | 16--18 | < one week | 6--9 |
| Rubella virus | near 0 | 14--18 | up to two weeks | largely asymptomatic, symptomatic period largely not infectious |
| Rhinovirus | near 0 | timecourse varies, may be infectious for weeks, may be wholly asymptomatic, may be symptomatic first day of infectivity. |  |  |
| Human respiratory syncytial virus | near 0 | may be wholly asymptomatic |  |  |
| Human parainfluenza | near 0 | may be wholly asymptomatic |  |  |
| Alphacoronavirus | near 0 | timecourse varies, may be wholly asymptomatic |  |  |

SARS-CoV-2 is modelled at a CFR of 1%, 1 week recovery period, and 3 days asymptomatic and infectious. Influenza is modelled at a CFR of 0.05%, 1 week recovery period, and 3 days asymptomatic and infectious. Smallpox infection is assumed to confer permanent immunity. SARS-CoV-2 and Influenza infections are assumed to confer immunity for one year on average. A constant birthrate of 2.5 births per 2 people over 100 years and death rate of one death per person over 100 years is assumed.

## Results

Endemic equilibrium is bounded within a range of (host) contact rates (Fig. 2B). When the mean time between contacts is too long and *k*_*I*_ is too low, 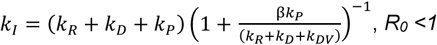, *R*_*0*_ <1, the pathogen goes extinct, and disease-free equilibrium is reached. Likewise, when contacts are too frequent, a bottleneck in the susceptible population occurs (Fig. 1B, here assumed to drop down to 10% of the total population), and the pathogen goes extinct. For some viruses, such as Smallpox and SARS-CoV-2, but not Influenza, a range of contact rates exists where the fraction of the infected population at endemic equilibrium is large enough so that the death rate exceeds the birth rate and the host-virus relationship is unsustainable in the long term, resulting in population decline without decreased pathogen virulence or modified host behavior. This range is very narrow for Smallpox but notably broad for SARS-CoV-2 (Fig. 2B, not shown to scale) encompassing a wider range of host behavior than endemic equilibrium. The existence of this region in the parameter space implies that herd immunity might be impossible to reach. In the middle-range of contact rates admitting endemic equilibrium (Fig. 1C), the decreased fraction of the infected population for all three examined viruses is offset by increasing CFR leading to an increased death rate. Under these model assumptions, the yearly death rate for SARS-CoV-2 is approximately 6 times that of Influenza.

To examine an expanded two-dimensional phase space, we allowed the CFR to vary from 10% to 100% for Smallpox-like viruses, with no asymptomatic spread and permanent immunity, and from 0% to 10%, for SARS-CoV-2-like and Influenza-like viruses, with asymptomatic spread and temporary immunity (Fig. 3). As the CFR increases, both threshold contact rates, corresponding to *R*_*0*_*=*1 and to the bottleneck in the susceptible population, increase and the range admitting endemic equilibrium narrows. Across much of the phase space, the host-pathogen relationship is unsustainable, and at very high CFR, the total host population falls below 10% (dark red) in the initial phase of the epidemic, signaling possible extinction at short timescales. The contours within the region corresponding to the endemic equilibrium indicate the total size of the infected population and are thus proportional to the size of the viral population and virus fitness. Within this region, increasing contact rate and decreasing CFR increases the size of the infected population. At extremely high CFR, the gradient points primarily in the direction of decreasing CFR, and at low contact rates, the gradient points primarily in the direction of increasing contact rate.

**Figure 3:**
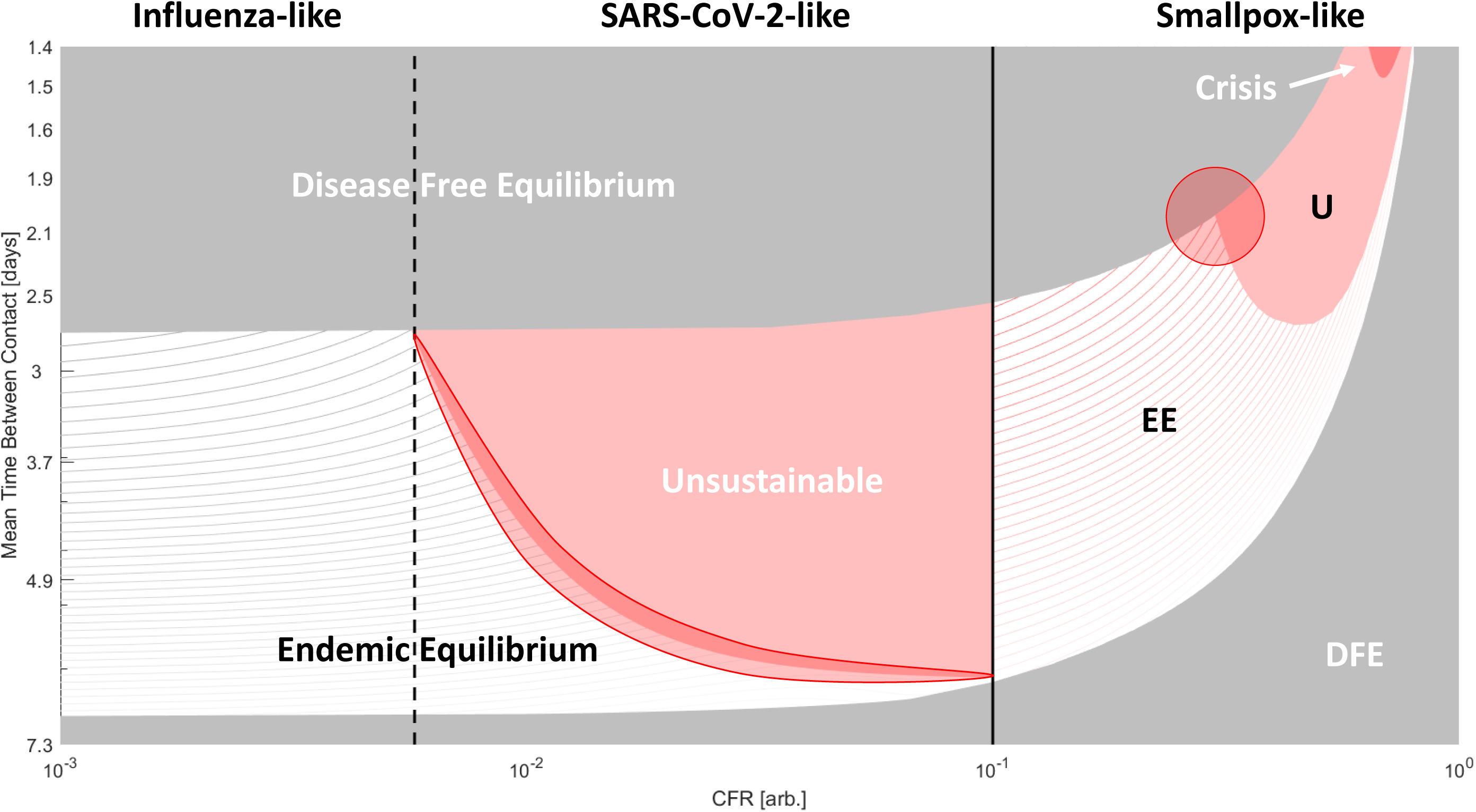
Phase diagram of epidemiological outcomes. The CFR and the contact rate were varied. Disease-free equilibrium(gray), endemic equilibrium(striped), host population decline(red), and host population crisis(loss of 90%, dark red) are shown. For CFR>10%, Smallpox-like features are assumed. For CFR<10% SARS-CoV-2/Influenza-like features are assumed (these viruses are only distinguished within the model by CFR). Lines within the region bounding endemic equilibrium indicate contours for the percentage of the population infected (*a*_*_ + *c*_*_)*100. Darker color corresponds to higher values. Two regions subject to kinetic constraints are highlighted.

Throughout most of the region in this phase space corresponding to endemic equilibrium for Smallpox-like viruses (Figure 3), moving Southwest increases the size of the infected population and suggests evolution towards decreased virulence. However, this is not the case at the Northeast corner representing viruses with extremely high CFR and extremely high contact rates (or infectivity). Such, hypothetical, viruses would have an unsustainable relationship with the host population if virulence decreased (and infected hosts were less likely to die before interacting with uninfected hosts) and are, in a sense, kinetically constrained, likely, by the host behavior. The population size of such viruses would dramatically increase if the CFR was reduced but would remain in endemic equilibrium only if contact rates or infectivity simultaneously decreased. Smallpox evolution could be similarly kinetically constrained. On the phase diagram for high CFR viruses, Smallpox, with a CFR of 30%, is located near the triple point for host populations with high contact rates where the regions of disease free equilibrium, endemic equilibrium and unsustainability meet (highlighted in Figure 3). To increase the size of the infected population, both CFR and contact rate must decrease, whereas decreasing only the CFR ultimately results in disease-free equilibrium when such a virus enters new host communities, due to a bottleneck in the size of the susceptible population.

The corresponding triple point located at the boundary between Influenza-like and SARS-CoV-2-like viruses lacks this property. For these viruses, decreasing CFR increases the size of the infected population throughout the parameter space. However, the boundary highlighted within the SARS-CoV-2 diagram (Fig. 3) represents a different kinetic constraint, this one, between immunity and mortality. While decreasing CFR below 10% minimally affects the threshold contact rates (*R*_*0*_*=*1 and susceptibility bottleneck), varying the duration of immunity has a dramatic impact on the phase diagram (Fig. 4A). As the duration of immunity decreases, with α = 0.01 corresponding to approximately 1.5 years at the phase boundary, the endemic equilibrium region shrinks substantially.

**Figure 4:**
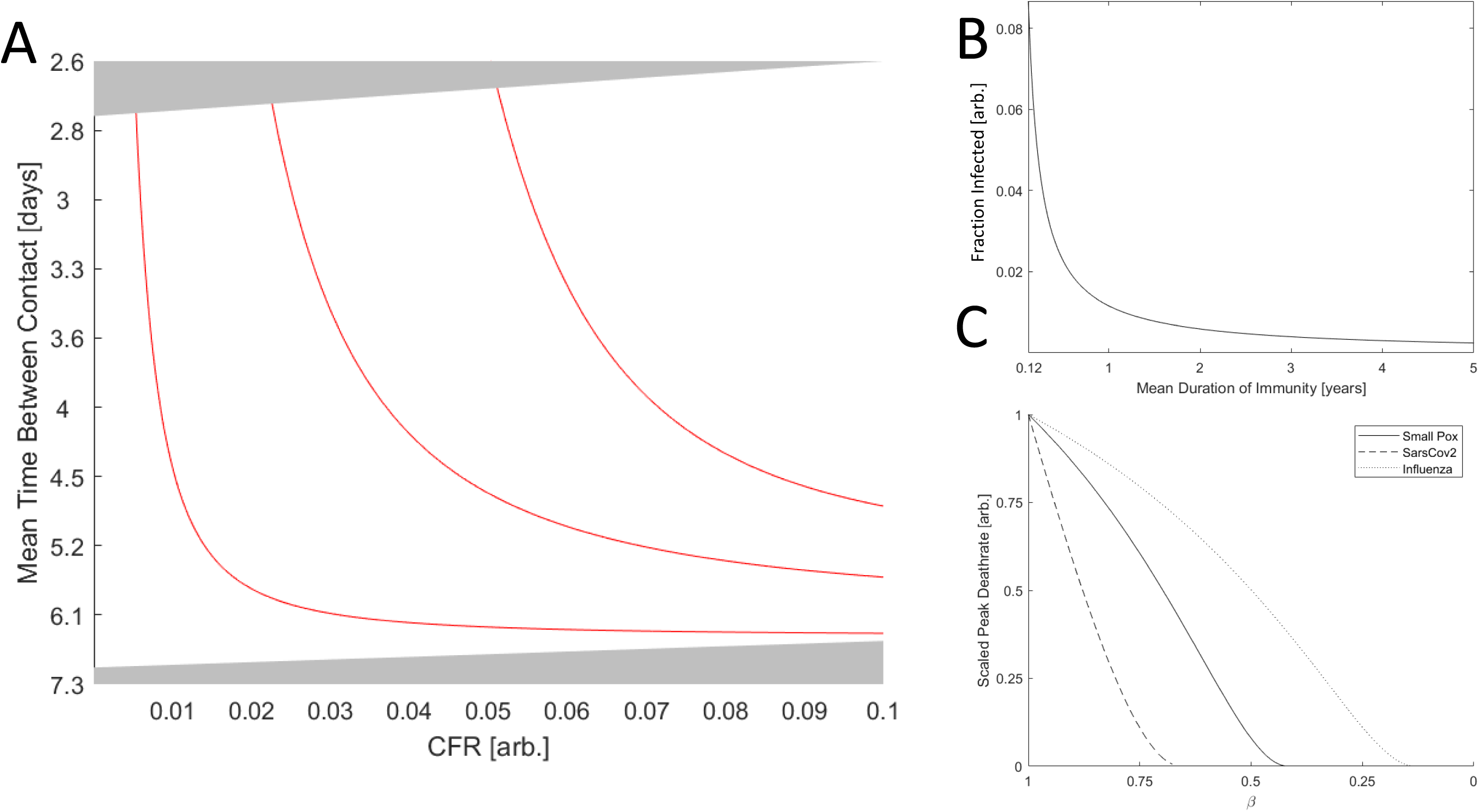
The role of immunity in epidemic evolution. **A**. Phase diagram of a SARS-CoV-2 like virus varying CFR and contact rate. Red lines indicate the boundary between endemic equilibrium and host population decline for α ∈ {0.01,0.045,0.08}. α = 0.01 corresponds to approximately 1.5 years of immunity. **B**. The fraction of the population infected, (*a*_*_ + *c*_*_), vs the duration of immunity for a hypothetical virus with a CFR of zero (other parameters matching Influenza and Sars-Cov-2) and the maximum admitted contact rate for endemic equilibrium. **C**. Peak death rate at the height of an epidemic with contact rate corresponding to the maximum value admitting endemic equilibrium given *β=0* and varying *β* (decreasing *β* corresponds to increasing isolation). Death rate is scaled by the maximum value for each virus.

As is apparent from the analytic solutions given in the Appendix (Additional File 1) and illustrated for a hypothetical virus with a CFR of zero (other parameters matching Influenza and Sars-Cov-2) and the maximum admitted contact rate (Fig. 4B), decreasing the duration of immunity dramatically increases the size of the infected population at endemic equilibrium across the parameter regimes. While typically viewed from a host-centric perspective, immunity is almost always necessary for the maintenance of endemic equilibrium and thus required for maintaining large viral populations over long time scales. Consider a SARS-CoV-2-like virus near the highlighted boundary in Figure 3. Suppose this virus acquires an adaptation enabling immunity evasion and thus decreasing the mean duration of immunity post infection. The size of the infected population will increase and, being near the boundary, the host-pathogen relationship will become unsustainable. This is another example of a kinetic constraint. Decreasing both CFR and the duration of immunity increases the size of the infected population, but maintaining a stable host-pathogen relationship and long term viral fitness requires that the reduction in CFR is proportional to the reduction in immune duration. Otherwise, the overall death rate for the host population might increase despite a reduction in pathogen virulence. Notably, decreasing the host-host contact rate moves the population farther from this boundary in the phase space and alleviates this kinetic constraint. Therefore, even if host-host contact rates cannot be reduced enough to break endemic equilibrium by reaching *R0*<1, a modest reduction can change the evolutionary trajectory of the pathogen from one of stagnant or increasing virulence to one of decreasing virulence.

One way a host population can effectively decrease the rate of infection is through the isolation of symptomatic individuals. Isolation decreases death rate at the peak of the epidemic (Fig. 4C) and can prevent an epidemic entirely by driving *R*_*0*_ below 1, which requires.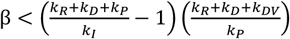 For pathogens with substantial asymptomatic or presymptomatic spread (small *k*_*P*_), this approach might not be feasible. Although isolation narrows the range of contact rates that admits endemic equilibria or unsustainability (Fig. 5A), the death rate at endemic equilibrium varies little with decreasing *β* and even increases in the case of SARS-CoV-2. Notably, however, SARS-CoV-2 is particularly sensitive to changes in *β* such that a modest decrease in *β*, that is, increased isolation, can change the long term outcome from endemic equilibrium to disease-free equilibrium. Furthermore, despite a 30-fold difference in CFR, at endemic equilibrium, Smallpox and SARS-CoV-2 have a comparable host mortality rate of approximately 0.1%/year, highlighting how pathogens with low or intermediate virulence can cause as many fatalities as highly virulent pathogens if allowed to reach endemic equilibrium.

**Figure 5:**
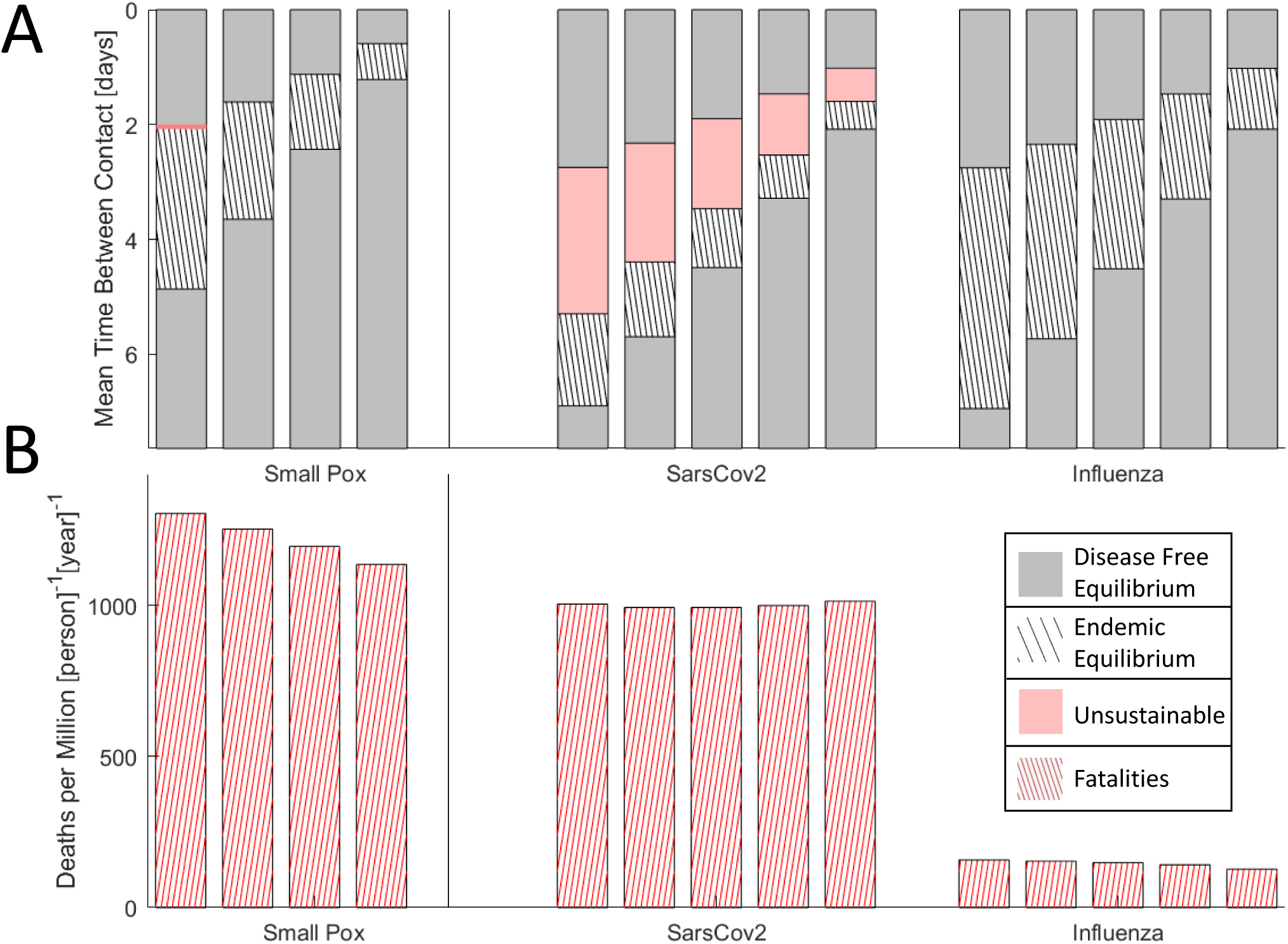
The effects of isolation on epidemic evolution. **A**. Disease free equilibrium (gray), endemic equilibrium (striped), and host population decline (red) for three viruses over a range of host contact rates. Red region for Smallpox is narrow and not shown to scale. Bars are ordered by decreasing β ∈ {0,0.25,0.5,0.75,1}. **B**. Death rate at endemic equilibrium (contact rates correspond to the midrange shown in A). Bars are ordered by decreasing β ∈ {0,0.25,0.5,0.75,1}.

## Discussion

We show here that virulence evolution in human pathogenic respiratory viruses can often be kinetically constrained. For Smallpox-like pathogens with high CFR in communities with frequent contact, a reduction in CFR can create a bottleneck in the size of the susceptible population, resulting in disease-free equilibrium (that is, extinction of the virus) when not accompanied by a decrease in the rate of infection. The rate of infection is determined by both the infectivity of the virus and the host-host contact rate. Under these conditions, if infectivity were internally constrained by CFR, this would not constitute a fitness trade-off and could facilitate host adaptation. On the other hand, high host-host contact rates could externally constrain the rate of infection making reduction in virulence costly to the virus. Evolution of the smallpox virus shows a steady pattern of gene losses that likely lead to increasing infectivity and virulence [30,31]. This evolutionary trend appears to be compatible with the conclusion that high CFR, Smallpox-like viruses are unlikely to evolve towards decreasing CFR due to constraints imposed by the host behavior.

For SARS-CoV-2-like pathogens with moderate CFR, evolution towards decreased virulence can be kinetically constrained by the relationship between CFR and the duration of immunity. Decreasing the duration of immunity increases the size of the infected population and the overall death rate which can make the host-pathogen relationship unsustainable. The existence of a large region of the phase space corresponding to unsustainable or kinetically constrained moderate CFR viruses implies two distinct forms of host response over two different timescales. Over long timescales, unsustainable viruses are likely to face extinction due to the elimination of the susceptible host population. Although, in principle, this could occur via extinction of the entire host population, the emergence of host resistance is likely. Over short timescales, especially for modern human populations, the emergence of an unsustainable virus, such SARS-CoV-2, may be considered societally unacceptable, leading to drastic measures that result in a major reduction in host-host contact rates. Both trends likely contribute to the paucity of moderate CFR human respiratory viruses which are subject to this kinetic constraint between immunity and virulence.

Perhaps paradoxically, immune evasion could incur a fitness cost for the pathogen and even lead to its extinction due to the host response. However, some level of immune evasion is required to maintain any state of endemic equilibrium in the case where lifelong immunity is conferred against individual strains and the duration of immunity is determined by antigenic drift rather than the decline of immunity itself. Evolution towards decreased virulence is uncertain in this case, and a better understanding of internal genomic constraints [7] could help predict the effects of immunomodulation. This is important when assessing the impact of novel or imperfect vaccination which can lead to counterintuitive results [32,33]. Diversification related to immune evasion commonly enables the maintenance of large virus populations over long time scales, as is the case for Influenza [34-36]. In the case of SARS-CoV-2, although immune evasion remains to be experimentally confirmed, diversification and host adaptation of SARS-CoV-2 have already been demonstrated [37-39]. Furthermore, products of virus genes that are specifically found in pathogenic beta-coronaviruses have been implicated in immunomodulation [40], suggesting that the virus adapts to maintain an endemic equilibrium in this way. Although the present model cannot predict whether SARS-CoV-2 will become more or less virulent, our results do suggest that its virulence evolution is kinetically constrained such that the region of the parameter space where reduced virulence could evolve is small. On the other hand, we show that even modestly decreasing the host-host contact rate can alleviate this kinetic constraint and promote virulence reduction.

During both the ongoing SARS-CoV-2 pandemic and the first Sars-Cov-1 epidemic, stringent public health measures were taken to limit transmission, extending beyond isolation of symptomatic individuals and into the quarantine of asymptomatic, and likely uninfected, contacts [17,25]. Although only a crude depiction of the nuanced dynamics underlying SARS-CoV-2 transmission, the analysis presented here suggests that isolation and quarantine are particularly effective towards changing the long-term outcome for viruses with moderate CFR and high infectivity, such as SARS-CoV-2. The phase diagram for SARS-CoV-2 is sensitive to the parameter *β* which reflects the isolation of symptomatic individuals. The evolution of the epidemic for such viruses is dominated by disease-free equilibrium or an unsustainable host-virus relationship. Endemic equilibrium is possible only in a narrow parameter range and is therefore unlikely. Nonetheless, the existence of this range suggests that herd immunity is unlikely and would amount to an extreme number of fatalities. Our analysis shows that, while highly amenable to public health intervention, without such efforts, SARS-CoV-2 can be expected to contribute to a substantially higher death toll than Influenza, comparable instead to that of Smallpox, for a protracted period.

## Conclusions

Human respiratory epidemics often evolve under kinetic constraints. These constraints can prevent the reduction in virulence for both high and moderate CFR viruses. The incorporation of these constraints can assist in the interpretation of classical model results for epidemics where some parameters, such as host-host contact rate, are unknown. We show that SARS-CoV-2 is unlikely to reach a state of endemic equilibrium; however, the potential for such equilibrium implies that herd immunity is likely unachievable. At equilibrium, moderate CFR viruses can cause as many fatalities as high CFR viruses with both SARS-CoV-2 and Smallpox leading to death of about 0.1% of the population per year. However, even partial isolation of symptomatic individuals can have a major effect not only by reducing the number of fatalities in the short term but also by potentially changing the evolutionary trajectory of the virus towards reduced virulence. Such simple public health interventions can dramatically decrease the forecasted cost of the virus over both the short and long term.

## Data Availability

NA

## Declarations

### Ethics approval and consent to participate: Not applicable. Consent for publication

Not applicable.

### Availability of data and materials

Not applicable.

### Authors’ contributions

NDR, YIW, and EVK conceived of and designed the study; NDR implemented the mathematical model; NDR, YIW, and EVK analyzed the results; NDR and EVK wrote the manuscript that was read and approved by all authors.

## Acknowledgements

The authors thank Koonin group members for helpful discussions.

## References

[1] Gandon S and Day T. The evolutionary epidemiology of vaccination. Journal of the Royal Society Interface, 2007;4(16):803–817.

[2] Lion S, and Gandon S. Spatial evolutionary epidemiology of spreading epidemics. Proceedings of the Royal Society B: Biological Sciences, 2016;283(1841):20161170.

[3] Frank SA. Models of parasite virulence. The Quarterly review of biology. 1996;71(1):37–78.

[4] Day T, Proulx SR. A general theory for the evolutionary dynamics of virulence. The American Naturalist. 2004;163(4):E40–63.

[5] Alizon S, de Roode JC, Michalakis Y. Multiple infections and the evolution of virulence. Ecology letters. 2013;16(4):556–67.

[6] Cressler CE, McLeod DV, Rozins C, Van Den Hoogen J, and Day T. The adaptive evolution of virulence: a review of theoretical predictions and empirical tests. Parasitology, 2016;143(7):915–930.

[7] Geoghegan JL and Holmes EC. The phylogenomics of evolving virus virulence. Nature Reviews Genetics, 2018;19(12):756–769.

[8] Alizon S, Hurford A, Mideo N, Van Baalen M. Virulence evolution and the trade-off hypothesis: history, current state of affairs and the future. Journal of evolutionary biology. 2009;22(2):245–59.

[9] Anderson RM, May RM. Coevolution of hosts and parasites. Parasitology. 1982;85(Pt 2):411–26.

[10] Gandon S, Mackinnon MJ, Nee S, Read AF. Imperfect vaccines and the evolution of pathogen virulence. Nature. 2001;414(6865):751–6.

[11] Mackinnon MJ, Read AF. Virulence in malaria: an evolutionary viewpoint. Philosophical Transactions of the Royal Society of London. Series B: Biological Sciences. 2004;359(1446):965–86.

[12] Soubeyrand B, Plotkin SA. Antitoxin vaccines and pathogen virulence. Nature. 2002;417(6889):609–10.

[13] Gandon S, Mackinnon MJ, Nee S, Read AF. Antitoxin vaccines and pathogen virulence. Nature. 2002;417(6889):610-.

[14] Lau LHL et al. Viral shedding and clinical illness in naturally acquired influenza virus infections. The Journal of infectious diseases, 2010;201(10):15091516.

[15] Bi Q et al. Epidemiology and transmission of covid-19 in 391 cases and 1286 of their close contacts in shenzhen, china: a retrospective cohort study. The Lancet Infectious Diseases, 2020.

[16] Lau EYH et al. A comparative epidemiologic analysis of sars in hong kong, beijing and taiwan. BMC infectious diseases, 2010;10(1):1–9.

[17] Barbisch D, Koenig KL, and Shih F. Is there a case for quarantine? perspectives from sars to ebola. Disaster medicine and public health preparedness, 2015;9(5):547–553.

[18] Jacobs SE, Lamson DM, St George K, and Walsh TJ. Human rhinoviruses. Clinical microbiology reviews, 2013;26(1):135–162.

[19] Munywoki PK et al. Frequent asymptomatic respiratory syncytial virus infections during an epidemic in a rural kenyan household cohort. The Journal of infectious diseases, 2015;212(11):1711–1718.

[20] CDC. https://www.cdc.gov/.

[21] UpToDate. https://www.uptodate.com/.

[22] Song Y et al. Asymptomatic middle east respiratory syndrome coronavirus infection using a serologic survey in korea. Epidemiology and health, 2018;40..

[23] Arino J et al. Simple models for containment of a pandemic. Journal of the Royal Society Interface, 2006;3(8):453–457.

[24] Brauer F. Some simple epidemic models. Mathematical Biosciences & Engineering, 2006;3(1):1.

[25] Lipsitch M et al. Transmission dynamics and control of severe acute respiratory syndrome. Science, 2003;300(5627):19661970.

[26] Diekmann O, Heesterbeek JAP, and Roberts MG. The construction of next-generation matrices for compartmental epidemic models. Journal of the Royal Society Interface, 2010;7(47):873–885.

[27] Guo H, Li MY, and Shuai Z. Global stability of the endemic equilibrium of multigroup sir epidemic models. Canadian applied mathematics quarterly, 2006;14(3):259–284.

[28] Sun R. Global stability of the endemic equilibrium of multigroup sir models with nonlinear incidence. Computers & Mathematics with Applications, 2010;60(8):2286–2291.

[29] Inc. The MathWorks. Symbolic Math Toolbox. Natick, Massachusetts, United State, 2019.

[30] Mühlemann B et al. Diverse variola virus (smallpox) strains were widespread in northern europe in the viking age. Science, 2020;369(6502).

[31] Hendrickson RC, Wang C, Hatcher EL, Lefkowitz EJ. Orthopoxvirus genome evolution: the role of gene loss. Viruses. 2010;2(9):1933–67.

[32] Soubeyrand B and Plotkin SA. Antitoxin vaccines and pathogen virulence. Nature, 2002;417(6889):609–610.

[33] Vale PF, Fenton A, and Brown SP. Limiting damage during infection: lessons from infection tolerance for novel therapeutics. PLoS Biol, 2014;12(1):e1001769.

[34] Wolf YI et al. Long intervals of stasis punctuated by bursts of positive selection in the seasonal evolution of influenza a virus. Biology direct, 2006;1(1):34.

[35] Gong LI, Suchard MA, Bloom JD. Stability-mediated epistasis constrains the evolution of an influenza protein. Elife 2, 2013;e00631.

[36] Kryazhimskiy S et al. Prevalence of epistasis in the evolution of influenza A surface proteins. PLoS Genet 7.2, 2011;e1001301.

[37] Rochman ND et al. Ongoing Global and Regional Adaptive Evolution of SARS3 CoV-2. bioRxiv. 2020.

[38] Y. Zhang et al. The ORF8 Protein of SARS-CoV-2 Mediates Immune Evasion through Potently Downregulating MHC-I. bioRxiv. 2020.

[39] Korber B et al. Tracking changes in SARS-CoV-2 Spike: evidence that D614G increases infectivity of the COVID-19 virus. Cell 2020;182.4, 812–827.

[40] Tan Y, Schneider T, Leong M, Aravind L, Zhang D. Novel Immunoglobulin Domain Proteins Provide Insights into Evolution and Pathogenesis of SARS-CoV-2-Related Viruses. Mbio. 2020;30;11(3).

