## Appendix for "Evolution of Human Respiratory Virus Epidemics"

Appendix A: derivation of $R_0$:

The linearized infection subsystem, in the limit of a completely susceptible population, is given by:

$$\begin{bmatrix} A \\ C \end{bmatrix}' = \begin{bmatrix} k_I - (k_R + k_D + k_P) & \beta k_I \\ k_P & -(k_R + k_D + k_{DV}) \end{bmatrix} \begin{bmatrix} A \\ C \end{bmatrix} \tag{1}$$

The rates, $R$, may be written as the sum of two matrices: $T$, transmission events leading to new infections and $E$, all other events:

$$T = \begin{bmatrix} k_I & \beta k_I \\ 0 & 0 \end{bmatrix}, E = \begin{bmatrix} -(k_R + k_D + k_P) & 0 \\ k_P & -(k_R + k_D + k_{DV}) \end{bmatrix} \tag{2}$$

The next generation matrix, $NGM$, may then be computed as the, sign corrected, product:

$$NGM = -TE^{-1} = \begin{bmatrix} \frac{k_I}{k_R+k_D+k_P} + \frac{\beta k_I k_P}{(k_R+k_D+k_P)(k_R+k_D+k_{DV})} & \frac{\beta k_I}{k_R+k_D+k_{DV}} \\ 0 & 0 \end{bmatrix} \tag{3}$$

with the characteristic equation:

$$-\lambda \left( \frac{k_I}{k_R + k_D + k_P} + \frac{\beta k_I k_P}{(k_R + k_D + k_P)(k_R + k_D + k_{DV})} - \lambda \right) = 0 \tag{4}$$

where $R_0$ is the dominant, in this case nonzero, eigenvalue.

Appendix B: solution for endemic equilibrium with constant population:

At endemic equilibrium the fraction of the total population, $N$, in each compartment, $X$, is constant: $\left(\frac{X}{N}\right)' = x' - xn' = x' = 0$ when the population is constant:

$$0 = \begin{bmatrix} i \\ s \\ a \\ c \end{bmatrix}' = \begin{bmatrix} -k_D & 0 & \alpha k_R & \alpha k_R \\ k_B & k_B - k_D - k_I \frac{a^*+\beta c^*}{i^*+s^*+a^*+\beta c^*} & k_B + (1-\alpha) k_R & k_B + (1-\alpha) k_R \\ 0 & k_I \frac{a^*+\beta c^*}{i^*+s^*+a^*+\beta c^*} & -(k_R + k_D + k_P) & 0 \\ 0 & 0 & k_P & -(k_R + k_D + k_{DV}) \end{bmatrix} \begin{bmatrix} i^* \\ s^* \\ a^* \\ c^* \end{bmatrix} \tag{5}$$

which asserts the additional constraint $k_B = k_D + k_{DV} c^*$. The epressions for $a^*$, $i^*$, and $s^*$ may be rewritten in terms of $c^*$. Subsequently enforcing unity, $i^* + s^* + a^* + c^* = 1$ yields a unique solution:

$$c^* = \left[ \alpha k_2 + \frac{k_3 (\alpha k_2 + k_1 + \beta)}{\left(1 + \frac{\beta}{k_1}\right) - k_3} + k_1 + 1 \right]^{-1} = \left[ \frac{R_0 - 1}{(\alpha k_2 + k_1 + 1) R_0 - (1 - \beta)} \right]^{-1} \tag{6}$$

where:

$$k_1 = \frac{k_R + k_D + k_{DV}}{k_P}, \; k_2 = \frac{k_R}{k_D}(1 + k_1), \; k_3 = \frac{k_R + k_D + k_P}{k_I} \tag{7}$$

As noted in the main text, this critical point does not correspond to a stable endemic equilibrium for all parameter regimes. For regimes where endemic equlibrium does exist, the derivative with respect to $\alpha/\beta$ can inform the impact of immunity and isolation of symptomatic hosts respectively:

$$\frac{\partial c^*}{\partial \alpha} = -(c^*)^{-2} \frac{k_R}{k_D} \left(1 + \frac{k_R + k_D + k_{DV}}{k_P}\right) \left[1 + \frac{1}{R_0 - 1}\right] \tag{8}$$

and

$$-\frac{\partial c^*}{\partial \beta} = -(c^*)^{-2} \left[ \frac{1}{(R_0 - 1)^2} \left(1 + \alpha \frac{k_2}{k_1 k_3}\right) \right] \tag{9}$$

One may additionally compare the limiting cases:

$$\frac{c^*(\alpha = 0)}{c^*(\alpha = 1)} = 1 + \frac{k_R}{k_D} \frac{R_0}{R_0 + \left(\frac{k_1+\beta}{k_1+1} - 1\right)} \geq 1 + \frac{k_R}{k_D} \tag{10}$$

which yields a decreasing function of $\beta$ and

$$\frac{c^*(\beta=1)}{c^*(\beta=0)} = 1+\frac{k_3}{1+\frac{1}{k_1}}\left(\frac{\left(1+\frac{1}{k_1}\right)-k_3}{1-k_3}\frac{(\alpha k_2+k_1)}{(\alpha k_2+k_1+1)}-1\right) \geq 1+\frac{k_1}{1-k_3}\left(\frac{k_3}{1+k_1}\right)^2 \tag{11}$$

which yields an increasing function of $\alpha$. Immunity and isolation work in concert.

Appendix C: general solution for endemic equilibrium:

All rates are normalized by the rate of infection.

$$0=\sum_{i=0}^{4}\left(\sum_j \lambda_{ij}k_{DV}^j\right)c^{*^i} \tag{12}$$

where:

$$\lambda_{43}=(1-\beta)\,k_P-1 \tag{13}$$

$$\lambda_{33}=2-(2-\beta)\,k_P \tag{14}$$

$$\lambda_{33}=\alpha k_R+\beta\left(k_P+3k_Bk_P+2k_Pk_R+k_P^2\right)+3k_B+k_P+2k_R-3k_Bk_P-2k_Pk_R-k_P^2 \tag{15}$$

$$\lambda_{23}=-2\alpha k_R+\beta\left(-k_P-2k_Bk_P-k_Pk_R-k_P^2\right)+5k_Bk_P-2k_P-2k_R-4k_B+3k_Pk_R+2k_P^2 \tag{16}$$

$$\lambda_{21}=-\alpha\beta k_Pk_R+\alpha\left(-2k_Bk_R-k_Pk_R-2k_R^2\right)+ \tag{17}$$

$$\beta\left(-3k_B^2k_P-2k_Bk_P^2-4k_Bk_Pk_R-2k_Bk_P-k_P^2k_R-k_P^2-k_Pk_R^2-k_Pk_R\right)+$$

$$3k_B^2k_P-3k_B^2+2k_Bk_P^2+4k_Bk_Pk_R-2k_Bk_P-4k_Bk_R+k_P^2k_R+k_Pk_R^2-k_Pk_R-k_R^2$$

$$\lambda_{12}=\alpha k_R+k_B+k_P-2k_Bk_P-k_Pk_R-k_P^2 \tag{18}$$

$$\lambda_{11}=\alpha\beta k_Pk_R+\alpha\left(2k_Bk_R+k_Pk_R+2k_R^2\right)+\beta\left(k_Bk_P+k_Bk_P^2+k_B^2k_P+k_P^2+k_Bk_Pk_R\right)+ \tag{19}$$

$$2k_B^2-4k_B^2k_P-3k_Bk_P^2-5k_Bk_Pk_R+3k_Bk_P+2k_Bk_R-k_P^2k_R-k_Pk_R^2+k_Pk_R$$

$$\lambda_{10}=\alpha\beta k_Pk_R\,(k_B+k_P+k_R)+\alpha\left(k_R^2+k_B*k_R\right)(k_B+k_P+k_R)+ \tag{20}$$

$$\beta\,(k_B+k_P+k_R)\left(k_Bk_P+k_B^2k_P+k_Bk_Pk_R\right)+(k_B+k_P+k_R)\left(k_Bk_R-k_B^2k_P+k_B^2-k_Bk_Pk_R\right)$$

$$\lambda_{01}=k_Bk_P\,(k_B+k_P+k_R-1) \tag{21}$$

$$\lambda_{00}=-\beta k_Bk_P^2+k_Bk_P\left(k_Bk_P-k_R-k_B+2k_Bk_R+k_Pk_R+k_B^2+k_R^2\right) \tag{22}$$

Appendix D: solution for endemic equilibrium in the limit of $c^* << 1$ and $k_P > k_R + k_{DV}$:

This limit yields the simplified system:

$$0=\begin{bmatrix} I/N \\ S/N \\ A/N \\ C/N \end{bmatrix}' \approx \begin{bmatrix} c^*k_{DV}-k_B & 0 & \alpha k_R & \alpha k_R \\ k_B & c^*k_{DV}-k_I\,(a^*+\beta c^*) & k_B+(1-\alpha)\,k_R & k_B+(1-\alpha)\,k_R \\ 0 & k_I\,(a^*+\beta c^*) & c^*k_{DV}-(k_R+k_P) & 0 \\ 0 & 0 & k_P & -(k_R+k_{DV}) \end{bmatrix}\begin{bmatrix} i^* \\ s^* \\ a^* \\ c^* \end{bmatrix} \tag{23}$$

First a note about $R_0$. In this case the linearized infection subsystem, in the limit of a completely susceptible population, becomes:

$$\begin{bmatrix} A/N \\ C/N \end{bmatrix}' \approx \begin{bmatrix} k_I - (k_R + k_P) + Ck_{DV} & \beta k_I \\ k_P & -(k_R + k_{DV}) \end{bmatrix} \begin{bmatrix} A/N \\ C/N \end{bmatrix} \tag{24}$$

We note this system is approximately the same as that introduced in Appendix A given $1 >> \frac{k_B, k_D}{k_R}$ and $1 >> c\frac{k_{DV}}{k_R}$ but may differ in general. The epressions for $a^*$ and $i^*$ may be rewritten in terms of $c^*$ while:

$$s^* \approx \frac{k_R + k_P}{k_I\left(1 + \frac{\beta}{\frac{k_R + k_{DV}}{k_P}}\right)} \approx \frac{1}{R_0} \tag{25}$$

yielding:

$$c^* \approx \frac{k_B}{k_{DV}}\left(1 + \alpha\frac{k_R\left(\frac{k_R + k_{DV}}{k_P} + 1\right)}{k_{DV}\left(1 - \frac{1}{R_0}\right)}\right)^{-1} \tag{26}$$

which represents a stable endemic equilibrium for sufficiently large alpha (at least partial immunity). The derivative with respect to $\beta$ can additionally inform the impact of isolation of symptomatic hosts:

$$-\frac{\partial c^*}{\partial \beta} \approx -\alpha\frac{k_I}{k_B}\frac{k_R + k_{DV} + k_P}{k_R + k_P}\frac{k_R}{k_R + k_{DV}}\left(\frac{c^*}{R_0 - 1}\right)^2 < -\alpha\frac{k_I}{k_B}\frac{k_R}{k_R + k_{DV}}\left(\frac{c^*}{R_0 - 1}\right)^2 \tag{27}$$

Appendix E: solution for endemic equilibrium in the limit of $1 >> \frac{k_{DV}}{k_R}$:

This limit yields the simplified system:

$$0 = \begin{bmatrix} I/N \\ S/N \\ A/N \\ C/N \end{bmatrix}' \approx \begin{bmatrix} c^*k_{DV} - k_B & 0 & \alpha k_R & \alpha k_R \\ k_B & c^*k_{DV} - k_I\frac{a^* + \beta c^*}{i^* + s^* + a^* + \beta c^*} & k_B + (1-\alpha)k_R & k_B + (1-\alpha)k_R \\ 0 & k_I\frac{a^* + \beta c^*}{i^* + s^* + a^* + \beta c^*} & -(k_R + k_P) & 0 \\ 0 & 0 & k_P & -k_R \end{bmatrix} \begin{bmatrix} i^* \\ s^* \\ a^* \\ c^* \end{bmatrix} \tag{28}$$

The epressions for $a^*$, $i^*$, and $s^*$ may be rewritten in terms of $c^*$ yielding:

$$i^* + s^* + a^* + c^* \approx \frac{\alpha k_R\left(\frac{k_R}{k_P} + 1\right)c^*}{k_B - c^*k_{DV}} + \frac{1 - (1-\beta)c^*}{R_0} + \left(\frac{k_R}{k_P} + 1\right)c^* = 1 \tag{29}$$

In the stricter limit $1 >> c^*\frac{k_{DV}}{k_B}$, this yields:

$$c^* \approx \frac{R_0 - 1}{\left(\alpha\frac{k_R}{k_B} + 1\right)\left(\frac{k_R}{k_P} + 1\right)R_0 + \beta - 1} \tag{30}$$

Appendix F: stability analysis:

The Jacobian matrix for the general case evaluated at the critical point is:

$$\boldsymbol{J^*} = \begin{bmatrix} -k_D & 0 & \alpha k_R & \alpha k_R \\ 0 & -\gamma k_I(a^* + \beta c^*) - k_D & -\gamma k_I s^* + (1-\alpha)k_R & -\gamma^2 k_I(\beta + (1-\beta)a^*)s^* + (1-\alpha)k_R \\ 0 & \gamma k_I(a^* + \beta c^*) & \gamma k_I s^* - (k_R + k_D + k_P) & \gamma^2 k_I(\beta + (1-\beta)a^*)s^* \\ 0 & 0 & k_P & -(k_R + k_D + k_{DV}) \end{bmatrix}, \gamma = (1 - (1-\beta)c^* \tag{31}$$

where the critical point is stable if all eigenvalues of the Jacobian are real and less than zero.
